# An Open-Label Multidose Psilocybin Intervention for Obsessive-Compulsive Disorder

**DOI:** 10.64898/2026.10.01.26364511

**Authors:** Praachi Tiwari, Sandeep M. Nayak, Nathan D. Sepeda, Rebecca Ehrenkranz, Michael A. Levine, Julia S. Rohde, Eliza Miller, Carina M. Beritela, Jeremy V. Scott, Gerald Nestadt, Frederick S. Barrett, David B. Yaden

## Abstract

**Importance:** Obsessive-Compulsive Disorder (OCD) has a 2% global prevalence, but only about 50% of patients respond to conventional treatment, underscoring the need for additional effective interventions.

**Objective:** To assess the feasibility, safety, and preliminary evidence for efficacy of repeated psilocybin administration in OCD patients who had previously attempted psychotherapy or pharmacotherapy.

**Design:** Waitlist-controlled open label randomized clinical trial.

**Setting:** Data were collected at the Johns Hopkins University School of Medicine.

**Participants:** Data were analyzed from 30 participants (total randomized N=37, by age, sex and OCD severity) with a failed previous attempt at either pharmacotherapy or evidence-based psychotherapies such as exposure response prevention (ERP).

**Intervention:** Participants received two psilocybin doses over two weeks (20 mg followed by 30 mg, if well-tolerated) under supportive conditions. The immediate-treatment arm received the drug within a month following enrollment, and the waitlist-control arm received identical support and dosing after an 8-week waiting period.

**Main Outcomes:** Clinician-administered Y-BOCS scores was collected one week post each drug administration and one month post session 2. Self-assessed reports were collected for acute subjective effects (Mystical Experience Questionnaire (MEQ), Challenging Experience Questionnaire (CEQ), and 11-Dimensional Altered States of Consciousness (11D-ASC)), State-Trait Anxiety Inventory (STAI), Beck Depression Inventory-II (BDI-II), and Quality of Life Enjoyment and Satisfaction Questionnaire (Q-LES-Q).

**Results:** A total of 37 participants were enrolled in the study (age: 38.4±11.6 years; 21 [56.75%] female), with 30 [81.08%] completing the 1-month follow-up. We show that two successive doses of psilocybin over two weeks were well-tolerated, leading to reduction in OCD symptoms. Compared with the waitlist group (N=14), the immediate-treatment group (N=16) showed a reduction in the Y-BOCS scores (F(2,56)=16.17, p<0.001, η^2^□=0.37) post session. This improvement persisted one month after the second psilocybin dose for both groups post treatment crossover for waitlisted group (F(3,84)=29.91, p<0.001, η^2^□=0.52). Y-BOCS scores were correlated with several acute subjective effects measures, including total scores on MEQ, and individual factors on CEQ and 11D-ASC. *Conclusions and Relevance:* Repeated psilocybin administration is tolerable and potentially efficacious in reducing OCD symptoms. These results support further investigation of multi dose psilocybin protocols as a viable therapeutic approach.

**Trial Registration:** ClinicalTrials.gov Identifier NCT05546658

**Key points:**

- Psilocybin was generally safe and tolerated across repeated dosing
- Psilocybin produced reductions in OCD symptom severity
- Quality of the psychedelic experience was correlated with therapeutic benefit

## Introduction

Obsessive-Compulsive Disorder (OCD) is a prevalent and debilitating mental health disorder, affecting approximately 1–2% of the global population^1–3^. OCD is characterized by intrusive thoughts (obsessions) and/or repetitive behaviors (compulsions) and can lead to significant distress and impairment in daily functioning^4,5^. Current treatment strategies primarily involve selective serotonin reuptake inhibitors (SSRIs) and cognitive-behavioral therapies such as exposure and response prevention (ERP)^4–13^. However, these approaches demonstrate limited population-wide efficacy, with only 40–60% of patients achieving substantial symptom reduction, highlighting a need for additional effective treatments^6,13–16^.

Classic psychedelic drugs have received increasing attention due to their potential long-lasting therapeutic effects for a growing range of psychiatric illnesses, especially mood and substance use disorders. Psilocybin has been widely studied in clinical trials for depression^17–20^ and substance use disorders^21–25^, with growing research in other mental health disorders. There has been limited investigation of psilocybin for OCD, and important questions remain regarding its safety, tolerability, dosing, and potential therapeutic effects in this population. Few clinical trials and a handful of case reports^26–30^ suggest that psychedelics might hold promise in alleviating OCD symptoms. The first clinical trial of psilocybin’s effects on OCD successfully assessed its acute safety and tolerability (N=9) with escalating doses (from low 100 μg/kg to moderately high 300 μg/kg, with a very low dose of 25 μg/kg inserted randomly after the first dose), each administered 1 week apart^31^. More recent clinical trials have also shown promising therapeutic effects of psilocybin in patients with OCD. A phase 1 study (N=19) assessing the effects of 1 mg oral psilocybin followed by a dose much lower than most clinical trials (10 mg, whereas most studies use 25 mg) showed reduction in compulsions (but not obsessions) for up to 1 week post intervention^32^. Yet another clinical trial with multiple exposures of different doses of psilocybin with at least four high dose interventions (300 ug/kg; N=5 per condition) over eight weeks was reportedly well-tolerated, and reduced OCD severity when compared to the active placebo (lorazepam) group^33^.

While emerging evidence suggests beneficial effects of psilocybin as pharmacotherapy for OCD, questions remain regarding optimal dosing (e.g. the utility of single vs multiple doses), appropriateness of comparators, and the role of concurrent pharmacological and/or behavioral therapy. Our work addresses some of these questions in a larger sample than previous studies and using sequential psilocybin doses, with conditional escalation from 20 mg (moderately high) to 30 mg (high) in participants with a previously failed attempt at conventional treatments. Given the open-label waitlist design, we primarily assessed safety endpoints and preliminary signals of therapeutic effects. Here, we provide evidence for the safety, tolerability, and potential efficacy of multiple escalating doses of psilocybin in patients with OCD.

## Materials and Methods

### Participants

Participants were recruited via online advertisements, paper flyers, and word of mouth. Interested individuals took an online pre-screener to assess initial eligibility before proceeding to phone screening by the study team. If participants were found provisionally eligible after pre-screening and phone screening, they signed consent and completed a comprehensive in-person eligibility assessment. Full eligibility criteria are reported in eMethods 1. A total of 37 participants with OCD who were otherwise medically and psychiatrically healthy were randomized (Figure 1), with the demographic distribution of participants as 62.2% White non-Hispanic, 16.2% White Hispanic, 5.4% White mixed population, 10.8% Asian mixed, and 5.4% Black. Out of these, 30 participants who completed all study visits were included in the complete-case analysis, with 16 participants identified as females, 12 as males, one as a transgender man, and one as non-binary. All participants provided their written informed consent prior to being enrolled in the study. The study was approved by an Institutional Review Board at the Johns Hopkins University School of Medicine and was performed in accordance with the 1964 Declaration of Helsinki.

**Figure 1:**
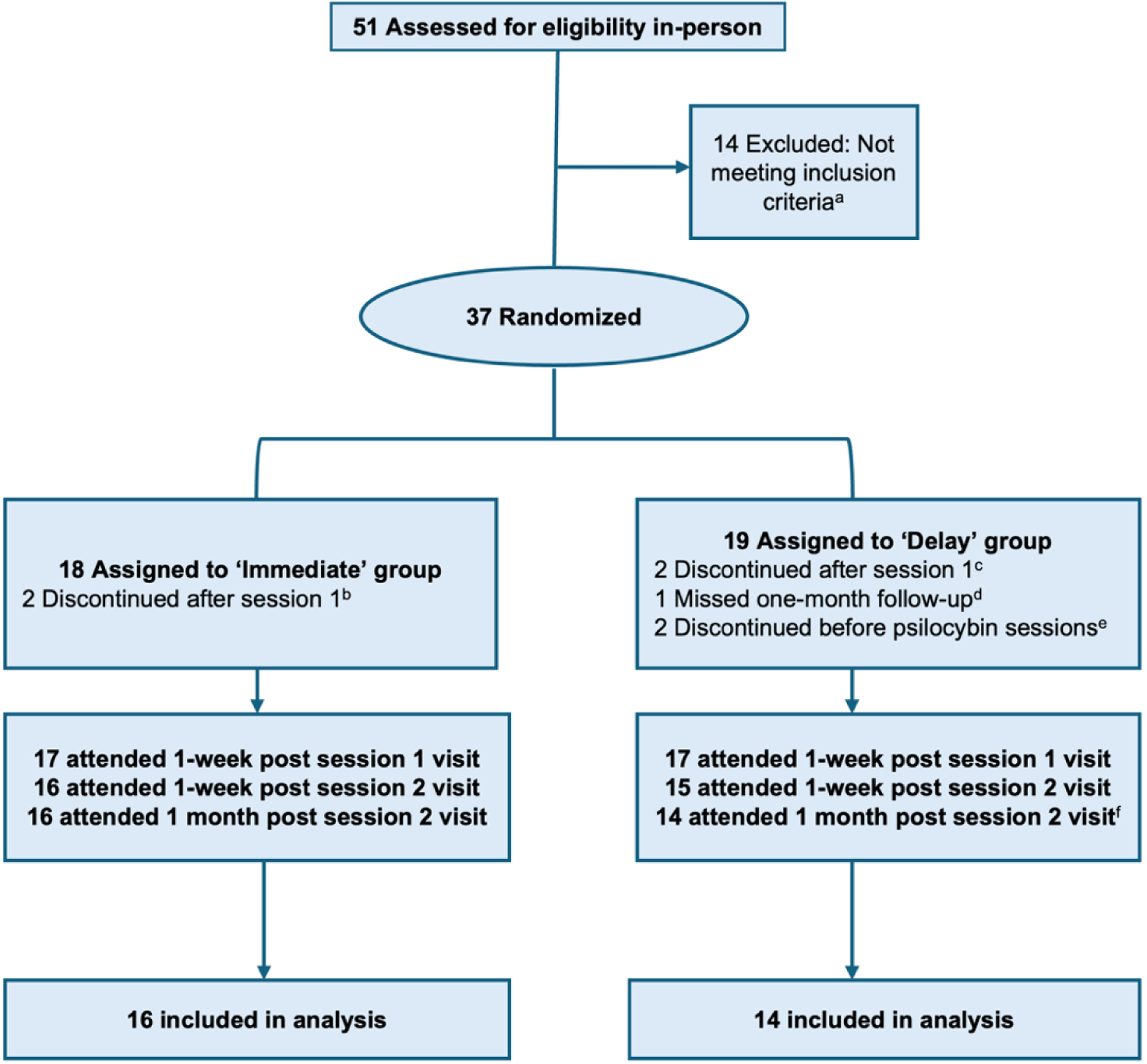
CONSORT flow diagram: 8156 online pre-screens and 284 phone screens were conducted. 51 participants meeting the initial inclusion/exclusion criteria were invited for in-person screening. ^a^ Reasons for the exclusion of 14 participants post in-person screening are detailed in eTable 1. ^b^ One participant withdrew due to unmet expectations from the first session, the second participant decided to restart their previous medication requiring them to withdraw. ^c^ Participants (2) withdrew after feelings of anxiety and/or panic after session 1. ^d^ Participant had scheduling conflicts which resulted in missing 1 month follow-up and all primary outcome measures. ^e^ One participant withdrew because they restarted therapy with improvement in OCD symptoms, the second participant had to withdraw because of sudden loss of a family member. ^f^ One participant misreported completion of self-assessed survey measures at one-month visit.

### Treatment paradigm

The study was designed as a randomized, waitlist-controlled trial to provide safety data and preliminary evidence for potential efficacy of psilocybin treatment for OCD. Participants were randomized using adaptive urn randomization^34^ balanced on sex, age and OCD symptom severity, to be assigned to either the ‘immediate’ or ‘delay’ group upon enrollment. Urn randomization was performed using *randPack* in R Statistical Software. If participants were on any medications that could render them ineligible (see inclusion/exclusion criteria), the participant tapered off of the medication for five half-lives prior to acceptance in treatment phase of study. Those assigned to the ‘immediate’ group began preparation for their study drug administration session immediately after enrollment (or after medication tapering, if needed), while the participants in the ‘delay’ group waited eight weeks after enrollment (in addition to the time required for medication taper, if needed) before they started preparatory sessions (Figure 2). All participants received psilocybin (from the Usona Institute). Each participant was assigned two study facilitators, at least one of whom was an independently licensed mental health professional with at least a master’s level of training in counseling or psychotherapy. Participants met with facilitators for preparation at least twice over the period of one month before the first dosing session. Preparation sessions consisted of discussing the trial logistics, providing psychoeducation on the potential effects of psilocybin, and ongoing assessment of safety issues, and the continued eligibility of the participants for the trial. The facilitators were present and available to support participants during the dosing sessions in cases of psychological distress as well as routine monitoring of blood pressure. The facilitators also met with participants for follow-up visits after the dosing sessions for reflective listening and evaluation of safety measures. Procedures followed on the drug administration day are detailed in eMethods 2.

**Figure 2:**
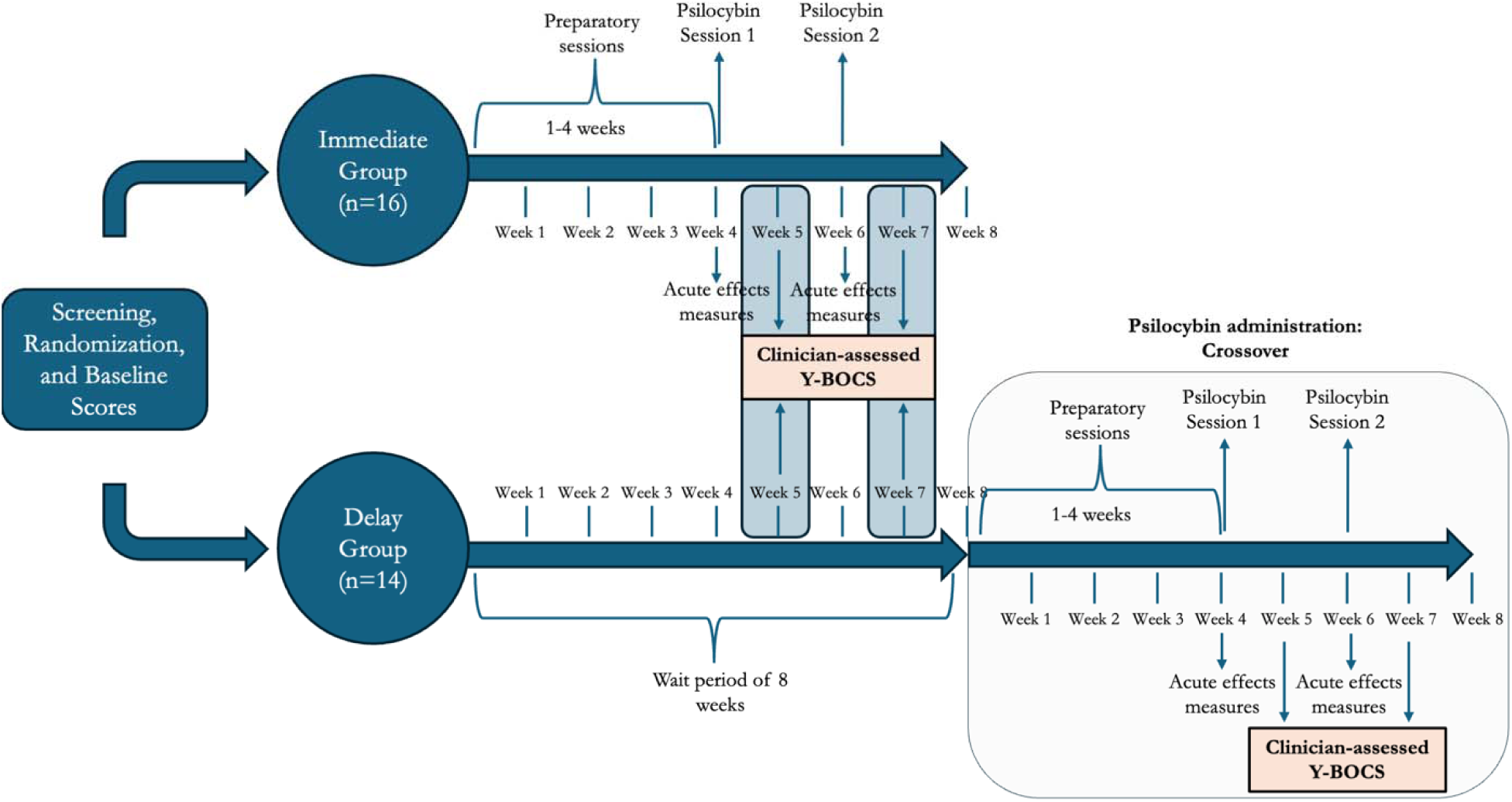
Schematic showing the trial design.

### Outcomes

The primary outcome of *Yale-Brown Obsessive-Compulsive Scale* (Y-BOCS) was assessed by the clinician (psychiatrist with forty years of clinical and research experience in OCD) to quantify OCD symptom severity^35,36^. Participants were also asked to complete self-assessment questionnaires for *Quality of Life Enjoyment and Satisfaction Questionnaire* (Q-LES-Q), depression using *Beck Depression Inventory II* (BDI-II), and anxiety using *State-Trait Anxiety Inventory* (STAI). These measures were measured one week after each dosing session and again one month after the second dosing session. Participants in the delayed group also completed these measures at weeks 5 and 7 after enrollment, corresponding to the 1-week post-session measurements being completed in the immediate group. The primary endpoints included change in clinician administered Y-BOCS score one-week post each intervention as compared to baseline, and change in BDI-II, STAI and Q-LES-Q scores, one month post second psilocybin dosing session from their respective baseline scores. Participants also completed self-assessed questionnaires associated with acute subjective effects (ASE) of psilocybin at the end of each dosing session, including the *Mystical Experience Questionnaire* (MEQ)^37,38^, the *Challenging Experience Questionnaire* (CEQ)^39^, and the *11-Dimensional Altered States of Consciousness Rating Scale* (11D-ASC)^40^. Additional information regarding measurement and outcomes is provided in eMethods 2. This study evaluates the primary analysis up to one month follow-up, and the 6-month data will follow in subsequent analysis and publications.

### Statistical Analysis

To evaluate the effects of psilocybin across the study timeline, we performed mixed-effects ANOVA models to assess Group, Time, and Group-by-Time effects, comparing the immediate and delayed treatment groups on all psychiatric outcome measures, Y-BOCS, BDI-II, STAI, and QLES-Q scores. Satterthwaite correction was applied if applicable. Post-hoc comparisons were performed, with the Bonferroni correction method applied to control for multiple comparisons. Effect sizes were estimated using partial eta-squared (η^2^□) for ANOVA effects and Cohen’s *d* for post-hoc pairwise comparisons. Pearson product-moment correlations were calculated to determine the association between acute subjective effects of psilocybin taken immediately after drug administration subsided and primary outcomes, as well as correlations among the primary outcome measures. False Discovery Rate correction via the Benjamini-Hochberg method was applied for multiple tests in Table 2. Statistical significance was defined as p<0.05 using a Neyman–Pearson framework. We assessed robustness to missing data from seven participants who withdrew after randomization using delta-adjusted tipping point analyses for the treatment-versus-waitlist and post-psilocybin comparisons (eMethods 2). All analyses were conducted in R (v4.3.1) using the *tidyverse* package suite^41^.

## Results

### 1. Psilocybin was generally well-tolerated and exhibited adequate safety profile in an OCD population

All participants enrolled in the study (N=37) reported limited adverse events and issues with tolerability. The most common of these were anxiety and headaches, with 29.41% and 61.36% of the respective reported incidents related to the study drug (Table 1). No unexpected or serious adverse events were reported during the study. Tolerability can be loosely inferred from study drop-outs and second dose decisions, the latter of which were based on tolerability of the first dose. A total of seven participants withdrew from the study (Figure 1). Out of the 30 participants included in the analysis, 24 participants (80%) received 30 mg for their second dose, and six participants (20%) proceeded with 20 mg as their second dose. We do not have sufficient evidence to determine whether a higher or lower dose was more effective; however, the higher dose was well tolerated and appeared relatively safe as part of the psilocybin dose-escalation procedure.

**Table 1:**
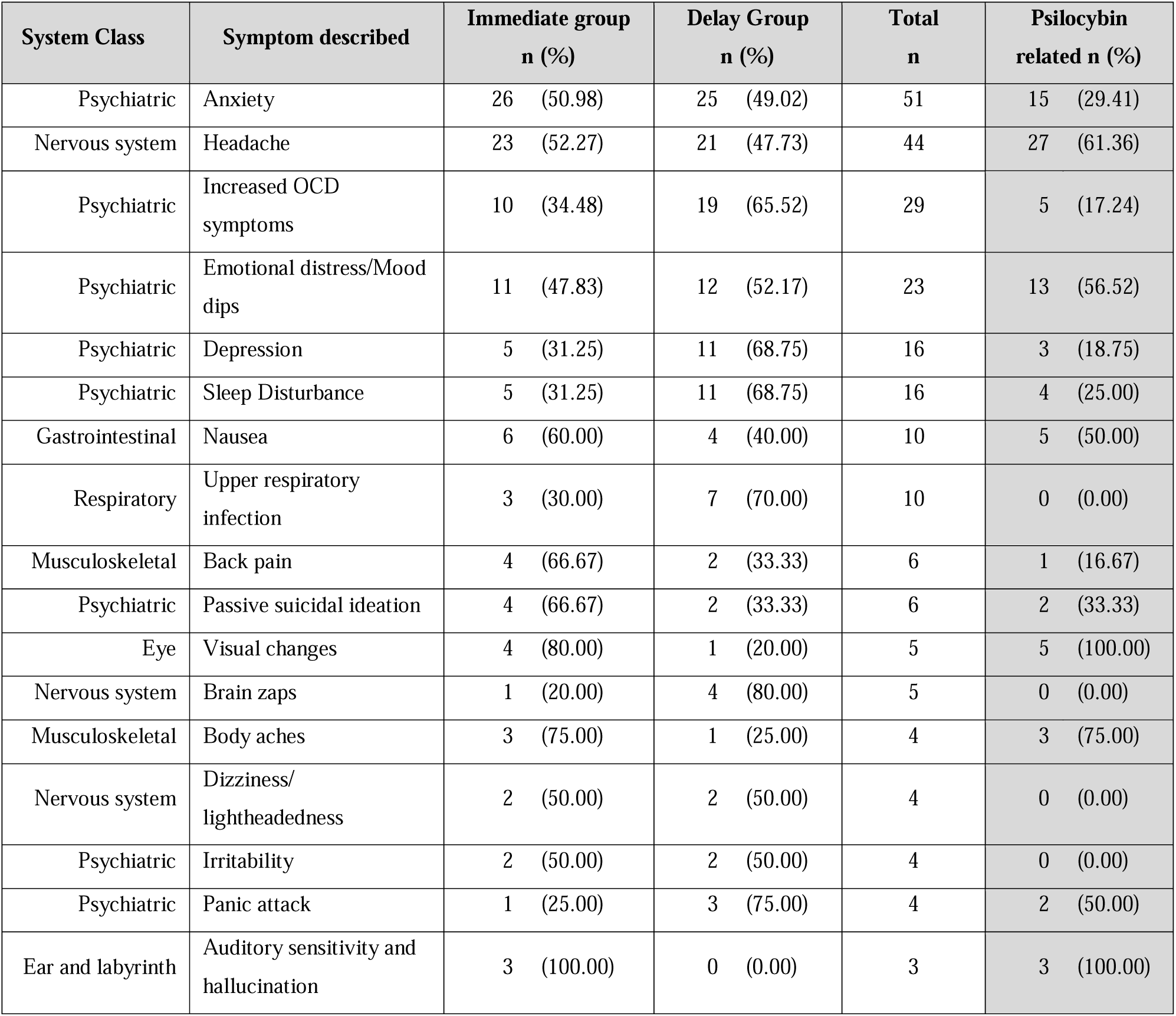
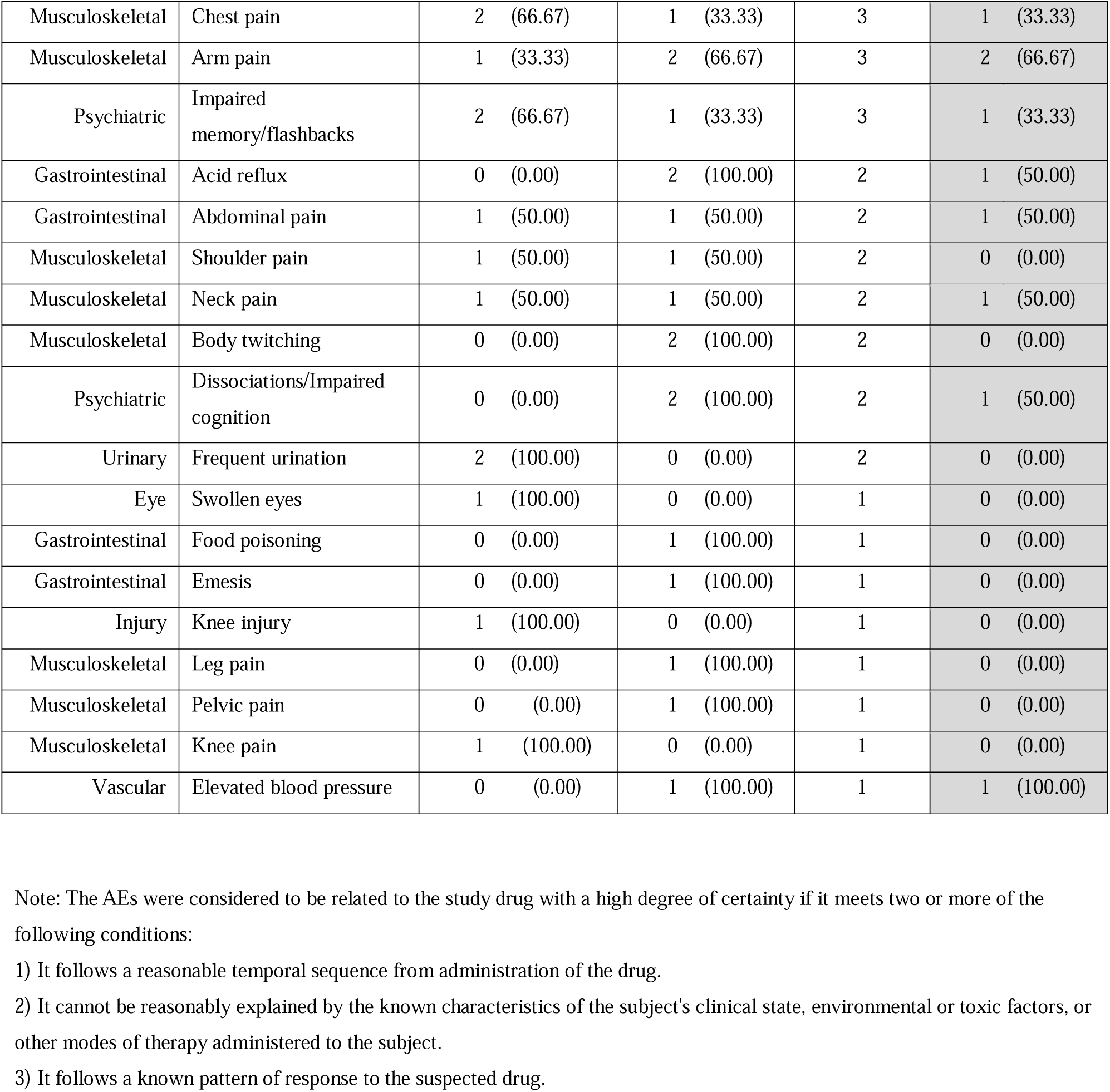
Summary of AEs as frequency of occurrence from enrollment up to one month post session 2.

### 2. Successive psilocybin doses result in reduction in OCD severity for both obsessions and compulsions

The between-group comparison on the primary outcome measure at the primary time-point was highly significant with a very large effect size. A comparison of clinician administered Y-BOCS scores between immediate (baseline: 25.9±4.3) and delay (baseline: 25.4±4.4) groups across the drug session timelines (Figure 3A) for drug vs no drug comparison showed a significant interaction between Group and Time (F(2,56)=16.17, p<0.001, η^2^□=0.37), with a main effect of both Group (F(1,28)=11.36, p=0.002, η^2^□=0.29) and Time (F(2,56)=15.51, p<0.001, η^2^□=0.36). Post-hoc analysis showed a significant decrease in average Y-BOCS scores from baseline to one week post session 1 (p=0.0049; Cohen’s d=1.21) and one week post session 2 (p<0.0001; Cohen’s d=1.92) in the immediate-treatment group (Y-BOCS score: post session 1= 17.25±7.96, post session 2: 13.44±9.47) as compared to the waitlist-control group (Y-BOCS score: delay week 5= 25±5.07, delay week 7: 25.71±4.55). In the immediate group, average Y-BOCS score also changed significantly after both first (p<0.0001; Cohen’s d=1.37) and second (p<0.0001; Cohen’s d=1.96) dosing sessions as compared to baseline. Y-BOCS scores decreased further between the assessments following the first and second sessions (p=0.005 Cohen’s d=0.60), although the study design does not permit attribution of this additional improvement specifically to the second psilocybin administration. Obsessions and compulsions, which are measured separately before being combined in the clinician administered Y-BOCS, were reduced to a roughly equivalent extent (Figure 3B, eMethods 3). We also compared clinician administered Y-BOCS scores before and after psilocybin treatment in all participants across both groups and noted a consistent decrease in average scores up to one month post session 2 (eFigure 1).

**Figure 3:**
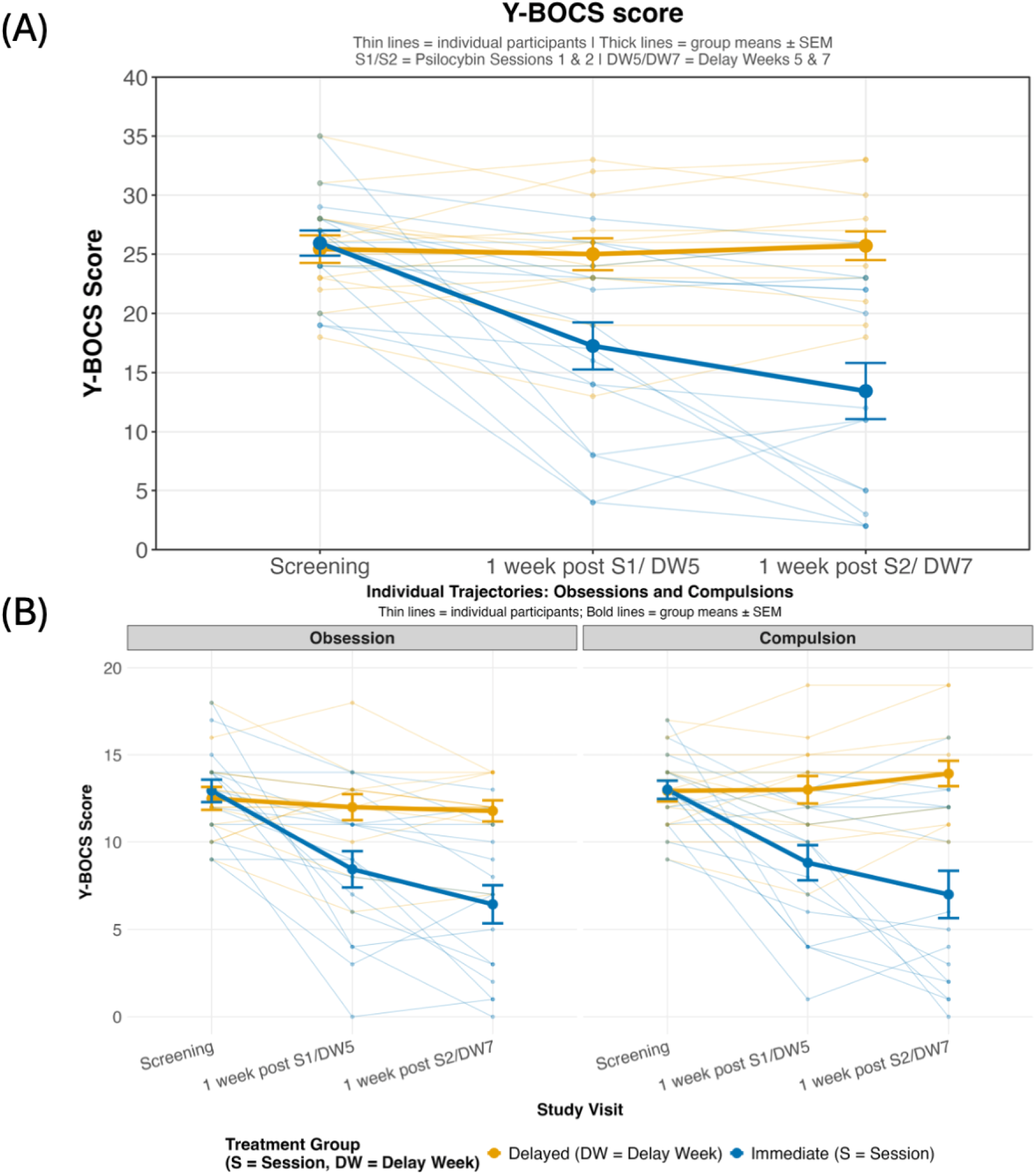
Successive psilocybin interventions reduce OCD severity for both obsessions and compulsions in the first 8 weeks post-enrollment. (A) Change in Y-BOCS score across two dosing sessions as compared between immediate (drug administered) and delay (no drug administered) groups. (B) Parcellation of obsession score and compulsion score across two dosing sessions as compared between immediate (drug administered) and delay (no drug administered) groups.

The reduction in clinician-administered Y-BOCS scores was substantial and clinically significant. A total of 40% of all participants after the first dosing session were responders (>35% from screening scores^42^) with 33.3% showing remission (Y-BOCS score≤12). After the second dosing session, 53.3% of total participants reached responder status with 40% showing remission. At one-month follow-up, 50% of all participants achieved responder status, with 36.7% in remission. Detailed group- and time-specific response of participants on Y-BOCS scores are elaborated in eTable 3. Our sensitivity analysis did not reveal any tipping point for our results that could be attributed to missing data (eFigure 3,4). Self-reported survey measures on STAI, BDI-II and Q-LES-Q were collected throughout the study timeline for all participants (eMethods 4, eFigures 4,5).

### 3. Some acute subjective effects are correlated with persisting YBOCS reductions

Some self-report measures of acute subjective effects (i.e., aspects of the psychedelic experience) collected after each session were correlated with OCD severity measured by the clinician administered Y-BOCS one-week post each session. Total scores on the MEQ, as well as factors including Bliss, Disembodiment, and Unity on the 11D-ASC, were negatively correlated (associated with improvements) with Y-BOCS score. While the total CEQ score was not significantly correlated with Y-BOCS scores, a significant positive correlation (associated with detriments) was observed for Y-BOCS score post psilocybin administration with Paranoia (Table 2).

**Table 2:** Correlation of measures of acute subjective effects to OCD score measured over two psilocybin sessions across two weeks. False Discovery Rate correction via the Benjamini-Hochberg method was applied for all tests. Data from both sessions was pooled, making n=60 for each correlated measure.

| Variable | Outcome | r | r squared | Cohen's d | p value |
| --- | --- | --- | --- | --- | --- |
| **Challenging Experience Questionnaire** |  |  |  |  |  |
| Death | Y-BOCS Score | -0.247 | 0.061 | -0.51 | 0.095 |
| Fear | Y-BOCS Score | 0.189 | 0.036 | 0.385 | 0.202 |
| Grief | Y-BOCS Score | 0.039 | 0.002 | 0.078 | 0.828 |
| Insanity | Y-BOCS Score | -0.029 | 0.001 | -0.058 | 0.868 |
| Isolation | Y-BOCS Score | 0.269 | 0.072 | 0.558 | 0.067 |
| Paranoia | Y-BOCS Score | 0.46 | 0.212 | 1.037 | 0.01 |
| Physical Distress | Y-BOCS Score | 0.124 | 0.015 | 0.249 | 0.411 |
| Total | Y-BOCS Score | 0.125 | 0.016 | 0.252 | 0.411 |
| **11-Dimensional Altered States of Consciousness** |  |  |  |  |  |
| Anxiety | Y-BOCS Score | 0.02 | 0 | 0.04 | 0.912 |
| Bliss | Y-BOCS Score | -0.36 | 0.13 | -0.772 | 0.013 |
| Changed perception | Y-BOCS Score | -0.198 | 0.039 | -0.405 | 0.199 |
| Complex imagery | Y-BOCS Score | -0.164 | 0.027 | -0.333 | 0.272 |
| Disembodiment | Y-BOCS Score | -0.45 | 0.203 | -1.009 | 0.002 |
| Elementary imagery | Y-BOCS Score | -0.175 | 0.031 | -0.355 | 0.241 |
| Impaired cognition | Y-BOCS Score | -0.041 | 0.002 | -0.082 | 0.828 |
| Insight | Y-BOCS Score | -0.282 | 0.08 | -0.588 | 0.056 |
| Spiritual | Y-BOCS Score | -0.285 | 0.081 | -0.594 | 0.055 |
| Synesthesia | Y-BOCS Score | -0.288 | 0.083 | -0.601 | 0.053 |
| Unity | Y-BOCS Score | -0.363 | 0.132 | -0.78 | 0.012 |
| **Mystical Experience Questionnaire** |  |  |  |  |  |
| Ineffability | Y-BOCS Score | -0.42 | 0.177 | -0.927 | 0.001 |
| Mystical | Y-BOCS Score | -0.45 | 0.202 | -1.007 | 0.002 |
| Positive | Y-BOCS Score | -0.486 | 0.236 | -1.112 | 0.001 |
| Transcendence | Y-BOCS Score | -0.431 | 0.186 | -0.955 | 0.002 |
| Total | Y-BOCS Score | -0.483 | 0.234 | -1.104 | 0.001 |

## Discussion

Our work highlights the safety and possible efficacy of a multi-dose psilocybin regimen, with up to two gradually escalating doses, for treating OCD in a patient population that frequently fails to respond to current conventional therapies. We also provide preliminary evidence that psilocybin reduces Y-BOCS scores to a clinically meaningful degree in OCD patients that persisted for at least one month after the second psilocybin intervention.

The results of our study were generally in line with results of prior studies that explored the safety, tolerability, and therapeutic potential of psilocybin across multiple neuropsychiatric disorders. The first clinical trial assessing psilocybin’s effects in OCD population was a double-blind randomized study that used four drug exposures ranging from sub-hallucinogenic to clearly hallucinogenic, and showed that while the Y-BOCS scores reduced within 24 hours after psilocybin, there was no significant persisting effect of dose (N=9)^31^. Scores returned to near baseline each week before the next intervention. In contrast, our study used two high doses in an open-label design in patients who had previously been treated with evidence-based psychotherapy or pharmacotherapy. We assessed clinician-administered Y-BOCS scores one week after each session and again up to one month after the second session to evaluate the sustained post-acute effects of psilocybin. We note that the general reduction in OCD severity persisted one month after the second dosing session. This timeline aligns with the results of another double-blind placebo-controlled trial (N=5/group), which used up to eight psilocybin interventions randomized between high (300 ug/kg) and low doses (100 ug/kg), with an active placebo control (lorazepam)^33^. This was followed by a cross-over of all participants to receive four high dose interventions in the second phase of the trial. The results show a significant reduction in OCD severity at the end of the eight-week treatment protocol, with 73.3% participants deemed responders (>35% reduction) and 40% as remitted (Y-BOCS score≤12)^33^. In our study, 50% of all participants achieved responder status, with 36.7% in remission at the one-month follow-up. Furthermore, we observe that both obsessions and compulsion reduce in severity one week after each psilocybin session. This is contrast with a previous study that a low-dose control (1 mg) for psilocybin followed by a 10 mg invention could reduce symptom severity on the Y-BOCS scale until one-week post administration, with a more promising effect on compulsions than obsessions. This study also shows that the reduction of Y-BOCS score from baseline in subsequent weeks is not statistically significant^32^, suggesting that future studies should test lasting effects with repeated low dose of 10mg psilocybin. This could also be a consequence of using one intervention of moderately low dose of psilocybin instead of a single high dose. Our study provides evidence that a persistent reduction in OCD severity could be possible with multiple high-dose interventions, although a longer-term impact of this intervention design remains to be evaluated. Overall, our work contributed to the growing body of literature that show the promising effects of psilocybin in treating OCD, in a variety of designs, repeated interventions, as well as variable persistence of effects.

## Limitations

The main limitation of our study is the absence of an active comparator. The open label study design allows us to infer safety and tolerability from the present study, with efficacy-related signals to be treated as preliminary evidence. All participants knew that the only drug they would be receiving was psilocybin, and they would eventually receive psilocybin despite the waiting period for the delay group. The obvious psychoactive effects of high-dose psilocybin has made it difficult to identify convincing active control conditions, although this work is ongoing. This design may therefore have amplified expectancy-related effects in participants receiving psilocybin^43^.

Moreover, the absence of improvement in the waitlist group should not be interpreted as evidence of a limited placebo response, because a waitlist control does not account for expectancy or other nonspecific effects associated with receiving the intervention. However, OCD by itself is known to have a low placebo response^44^. Furthermore, it is extremely important that future studies in this population include an active comparator that can distinguish the effects of the study drug compared to a comparator in lieu of a placebo response.

Another limitation was that the study required the participants to taper off medications. While we cannot know for sure, this may have impacted the Y-BOCS scores of participants in the delayed group showing less improvement than the immediate treatment group at one month post session 2, reflecting aspects of the delayed group’s treatment sequence in which medication tapering required by some participants was followed by a wait period that may have resulted in a longer interval without pharmacotherapy before psilocybin administration, as well as expectations of a psychedelic experience or disappointment related to the wait period.

## Conclusion

In this small, open-label randomized trial, two administrations of psilocybin were generally well tolerated in adults with OCD who had previously received evidence-based treatment. Immediate treatment was associated with greater reductions in OCD symptoms than waiting, providing preliminary evidence that repeated psilocybin administration warrants further investigation in adequately powered, controlled trials. Greater acute subjective effects on all MEQ subscales and on the Bliss, Unity, and Disembodiment subscales of the 11D-ASC were associated with greater reductions in Y-BOCS scores, whereas greater Paranoia on the CEQ were associated with smaller reductions in Y-BOCS scores. Longer-term follow-up, including assessments 6 months or later, is needed to determine the durability of these effects. Most importantly, this study should be followed by a fully powered RCT with an appropriate active control condition.

## Data Availability

All data produced in the present study are available upon reasonable request to the authors.

## eAppendices

### Supplement Online content

#### eMethods 1: Eligibility Criteria

<u>Ages Eligible for Study:</u> 21 Years to 80 Years

<u>Genders Eligible for Study:</u> All

<u>Accepts Healthy Volunteers:</u> No

##### Inclusion Criteria

1. Have given written informed consent
2. Currently meet criteria for a DSM-5 diagnosis of OCD and report a history of OCD for at least 1 year prior to screening
3. Have a Y-BOCS score of 18 or more
4. Have at least one prior attempt at treatment, either ERP or pharmacotherapy
5. No antidepressant medications for approximately five half-lives prior to acceptance in treatment phase of study
6. Women who are of childbearing potential and sexually active who are not practicing an effective means of birth control must agree to practice an effective means of birth control throughout the duration of the study
7. Be judged by study team clinicians to be at low risk for suicidality
8. Concurrent psychotherapy is allowed if the type and frequency of the therapy has been stable for at least two months prior to screening and is expected to remain stable during participation in the study
9. Be otherwise medically stable as determined by screening for medical problems via a personal interview, a medical questionnaire, a physical examination, an electrocardiogram (ECG), and routine medical blood and urinalysis laboratory tests (CBC, CMP, urine beta-HCG, urine toxicology screen)
10. Agree to consume approximately the same amount of caffeine-containing beverage (e.g., coffee, tea) that he/she consumes on a usual morning, before arriving at the research unit on the mornings of drug session days. If the participant does not routinely consume caffeinated beverages, he/she must agree not to do so on session days
11. Agree to refrain from using any psychoactive drugs, including alcoholic beverages, within 24 hours of each drug administration. The exceptions are caffeine and nicotine
12. Agree not to take any PRN medications on the mornings of drug sessions
13. Agree not to take sildenafil (Viagra®), tadalafil, or similar medications within 72 hours of each drug administration
14. Agree that for one week before each drug session, he/she will refrain from taking any nonprescription medication, nutritional supplement, or herbal supplement except when approved by the study investigators. Exceptions will be evaluated by the study investigators and will include acetaminophen, non-steroidal anti-inflammatory drugs, and common doses of vitamins and minerals.
15. Have limited lifetime use of hallucinogens (the following criteria are preferred: no use in the past 5 years; total hallucinogen use less than 10 times)

##### Exclusion Criteria

1. Clinically significant transaminitis (AST or ALT greater than two times normal value)
2. Women who are pregnant (as indicated by a positive urine pregnancy test assessed at intake and before each drug session) or nursing
3. Women who are of childbearing potential and sexually active who are not practicing an effective means of birth control
4. Cardiovascular conditions: coronary artery disease, stroke, angina, uncontrolled hypertension, a clinically significant ECG abnormality (e.g., atrial fibrillation), prolonged QTc interval (i.e., QTc > 450 msec), heart valve, or transient ischemic attack (TIA) in the past year
5. Epilepsy with history of seizures
6. Type 1 diabetes
7. BMI < 18
8. Currently taking on a regular (e.g., daily) basis, any psychoactive prescription medication or any medications having a primary centrally acting serotonergic effect, or MAOIs. For individuals who have intermittent or PRN use of such medications, psilocybin sessions will not be conducted until approximately five half-lives of the agent have elapsed after the last dose.
9. Current (severe) migraine or other recurring severe headaches
10. Current or past history of meeting DSM-5 criteria for schizophrenia spectrum or other psychotic disorders (except substance-/medication-induced or due to another medical condition), or bipolar I disorder
11. Current or history within one year of meeting DSM-5 criteria for a moderate or severe alcohol, or other drug use disorder (excluding tobacco and caffeine)
12. Nicotine dependence that would be incompatible with an individual to be nicotine free for 8-10 hours on a psilocybin session day
13. Have a first degree relative with schizophrenia or other psychotic disorders (except substance/medication-induced or due to another medical condition), or bipolar I disorder

#### eMethods 2: Methods Description

Procedures followed on the session days:

On drug administration days, participants were assessed for normotension and normocardia, absence of pregnancy (for females), and use of illicit substances before proceeding with dosing. During drug administration, participants were asked to lay down on the couch wearing eyeshades and earphones playing music. Participants were asked to listen to a standardized playlist of songs that was preselected by the study team, featuring music intended to provide a comfortable background sound. Blood pressure and heart rate were monitored every 30 minutes for the first two hours and then once every hour from the time of drug administration for up to six hours. A physician was on call and available to respond to medical or psychiatric issues for the duration of the drug administration session, and rescue medications (eTable 2) were available to treat hypertensive crisis or adverse subjective experience, if needed. Participants were monitored until the acute effects of psilocybin had subsided and were required to be medically and psychologically stable before leaving the study site. On the day of the first session, participants received 20 mg of oral psilocybin, administered with water. If the initial dose was well tolerated as per subjective assessment by the study team, a higher dose of 30 mg would be used in the second dosing session conducted approximately two weeks later. In cases where the first session was associated with psychologically or physiologically challenging effects, the second session proceeded with 20 mg.

##### Tipping point sensitivity analysis

To evaluate the robustness of our findings to missing data from the seven participants who withdrew after randomization, we conducted tipping point sensitivity analyses for both the between-group treatment-versus-waitlist comparison as well as the post-psilocybin trajectory comparison. For each analysis, missing Y-BOCS scores were imputed using the mean of completers within the same treatment group, adjusted by a delta parameter representing a change in Y-BOCS scores. A delta of 0 assumes equivalence between dropouts and completers, and positive delta values assume worse outcomes (e.g., a delta of 5 assumes dropouts scored 5 points higher on the Y-BOCS than completers). The original mixed-effects model was refit for each delta value to identify the tipping point, defined as the delta at which the Group x Time interaction changes. For the psilocybin versus waitlist control comparison, both withdrawn participants belonged to the immediate treatment group. Because the threat to validity is that treated dropouts may have responded less favorably than completers, delta was applied in the positive direction only, ranging from 0 to 27, at which imputed Y-BOCS scores would exceed the ceiling of 40. For the post-psilocybin comparison, both groups had received psilocybin and there was no directional expectation for how dropouts would differ from completers. Delta ranged from −19 to +28, spanning the full range of possible Y-BOCS scores. The tipping point was defined as the delta value at which the Group x Time interaction crossed the α = 0.05 threshold.

##### Data Scoring

*i. Yale-Brown Obsessive-Compulsive Scale (Y-BOCS) score*: There are ten Y-BOCS items, and each item is scored on a five-point scale from 0 = "no symptoms" to 4 = "extreme symptoms". The sum of the first five items is a severity index for obsessions, and the sum of the last five an index for compulsions. Total score is used as a measure of OCD severity. The scale ranges from 0-40, with 0-20 for obsessions and 0-20 for compulsions.
*ii. Beck Depression Inventory II (BDI-II):* The BDI-II is a self-report inventory of depressive symptoms, consisting of 21-items. Each of the 21 items corresponding to a symptom of depression is summed to give a single score for the BDI-II. There is a four-point scale for each item ranging from 0 to 3. On two items (16 and 18) there are seven options to indicate either an increase or decrease of appetite and sleep. Higher scores indicate higher levels of depression.
*iii. State-Trait Anxiety Inventory (STAI):* The State-Trait Anxiety Inventory has 40 items on a 4-point scale, with each subscale containing 20 questions each (State and Trait). Responses for the State anxiety scale assess intensity of current feelings "at this moment", while responses for Trait anxiety scale assess intensity of long standing, persistent feeling with following scores: 1) not at all, 2) somewhat, 3) moderately so, and 4) very much so. Item scores are added to obtain subtest total scores. Scoring is reversed for anxiety-absent items. Range of scores for each subtest is 20-80, with higher scores indicating greater anxiety.
*iv. Quality of Life Enjoyment and Satisfaction Questionnaire (Q-LES-Q):* The Quality of Life Enjoyment and Satisfaction Questionnaire - Short Form (Q-LES-Q-SF) is a 16-item self-report questionnaire that captures life satisfaction over the past week. Each question is scored on a 5-point scale from 1 (Very Poor) to 5 (Very Good) that indicates the degree of enjoyment or satisfaction achieved during the past week relative to the particular activity or feeling described in the item. Higher score indicates higher quality of life.

**eTable 1:**
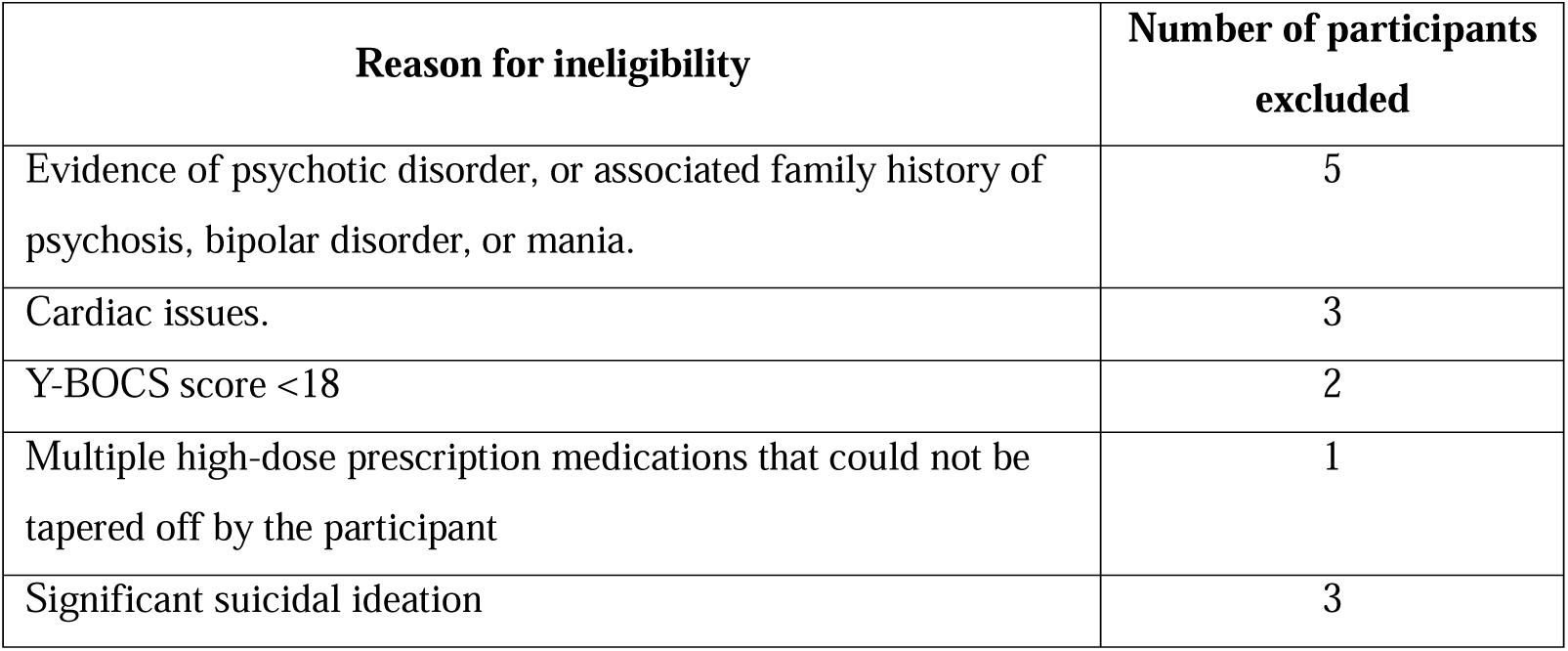
Reason for ineligibility of the 14 participants excluded from the study after in-person screening.

**eTable 2:** Rescue medications used in the study.

| Indication | Rescue medication | Dose | Usage Frequency | Reason |
| --- | --- | --- | --- | --- |
| Acute anxiety, agitation, or panic | Diazepam (oral) | 5–10 mg | 3 | Panic, anxiety |
| Acute psychosis | Risperidone (oral or orally disintegrating tablet) | 0.5–1 mg | 1 | Panic due to acute effects |
| Acute nausea/emesis | Ondansetron (oral) | 4–8 mg | 1 | Nausea |
| Hypertensive emergency | Captopril (oral) | 25 mg | - | - |
| Refractory hypertension (if captopril is insufficient) | Clonidine (oral) | 0.1 mg | - | - |

**eTable 3:** Summary of change in Y-BOCS scores and participants in remission at each timepoint.

|  |  | Percent average<br>change in Y-BOCS score<br>(% change) | Percentage of<br>participants with<br>responder status<br>(>35%) | Percentage of<br>participants in<br>remission<br>(Y-BOCS≤12) |
| --- | --- | --- | --- | --- |
| **Medication tapering (n=11)** |  |  |  |  |
| *Immediate group*<br>(n=4) | One week post session 1 | -21.22 | 25.00 | 25.00 |
|  | One week post session 2 | -28.32 | 25.00 | 25.00 |
|  | One month post session 2 | -56.38 | 50.00 | 50.00 |
| *Delayed group*<br>(n=7) | One week post session 1 | -27.49 | 42.86 | 28.57 |
|  | One week post session 2 | -16.38 | 28.57 | 14.29 |
|  | One month post session 2 | -15.03 | 28.57 | 14.29 |
| **No medication tapering (n=19)** |  |  |  |  |
| *Immediate group*<br>(n=12) | One week post session 1 | -37.02 | 41.67 | 25.00 |
|  | One week post session 2 | -55.18 | 66.67 | 66.67 |
|  | One month post session 2 | -52.44 | 58.33 | 50.00 |
| *Delayed group*<br>(n=7) | One week post session 1 | -46.58 | 42.86 | 57.14 |
|  | One week post session 2 | -44.63 | 71.43 | 28.57 |
|  | One month post session 2 | -32.78 | 57.14 | 28.57 |
| **Total (n=30)** |  |  |  |  |
| *Immediate group*<br>(n=16) | One week post session 1 | -33.07 | 37.50 | 25.00 |
|  | One week post session 2 | -48.46 | 56.25 | 56.25 |
|  | One month post session 2 | -53.42 | 56.25 | 50.00 |
| *Delayed group*<br>(n=14) | One week post session 1 | -37.04 | 42.86 | 42.86 |
|  | One week post session 2 | -30.51 | 50.00 | 21.43 |
|  | One month post session 2 | -23.91 | 42.86 | 21.43 |

#### eMethods 3: Y-BOCS scores separated by Obsession and Compulsion components were compared between immediate and waitlist-control arm

A 3-way mixed-effects ANOVA showed no significant interaction of Group x Time x Measure (obsession vs compulsion subscales of the Y-BOCS; F(2,140)=0.29, p=0.750, η^2^□=0.004) but revealed a significant interaction for Group x Time (F(2,140)=29.59, p<0.001, η^2^□=0.30).

A 2-way analysis for the obsession sub-scale revealed a significant interaction for Group x Time (F(2,56)=12.01, p<0.001, η^2^□=0.30), with a main effect of both Group (F(1,28)=8.61, p=0.007, η^2^□=0.24) and Time (F(2,56)=18.69, p<0.001, η^2^□=0.40). Post-hoc analysis showed a significant difference between delay and immediate groups at one week post session 1 (p=0.012; Cohen’s d=1.10) as well at one week post session 2 (p<0.001; Cohen’s d=1.65).

Similarly for the compulsion sub-scale, a 2-way analysis revealed a significant interaction for Group x Time (F(2,56)=15.88, p<0.001, η^2^□=0.36), with a main effect of both Group (F(1,28)=12.20, p=0.002, η^2^□=0.30) and Time (F(2,56)=9.09, p<0.001, η^2^□=0.25). Post-hoc analysis showed a significant difference between delay and immediate groups at one week post session 1 (p=0.005; Cohen’s d=1.20) as well at one week post session 2 (p<0.001; Cohen’s d=1.98).

**eFigure 1:**
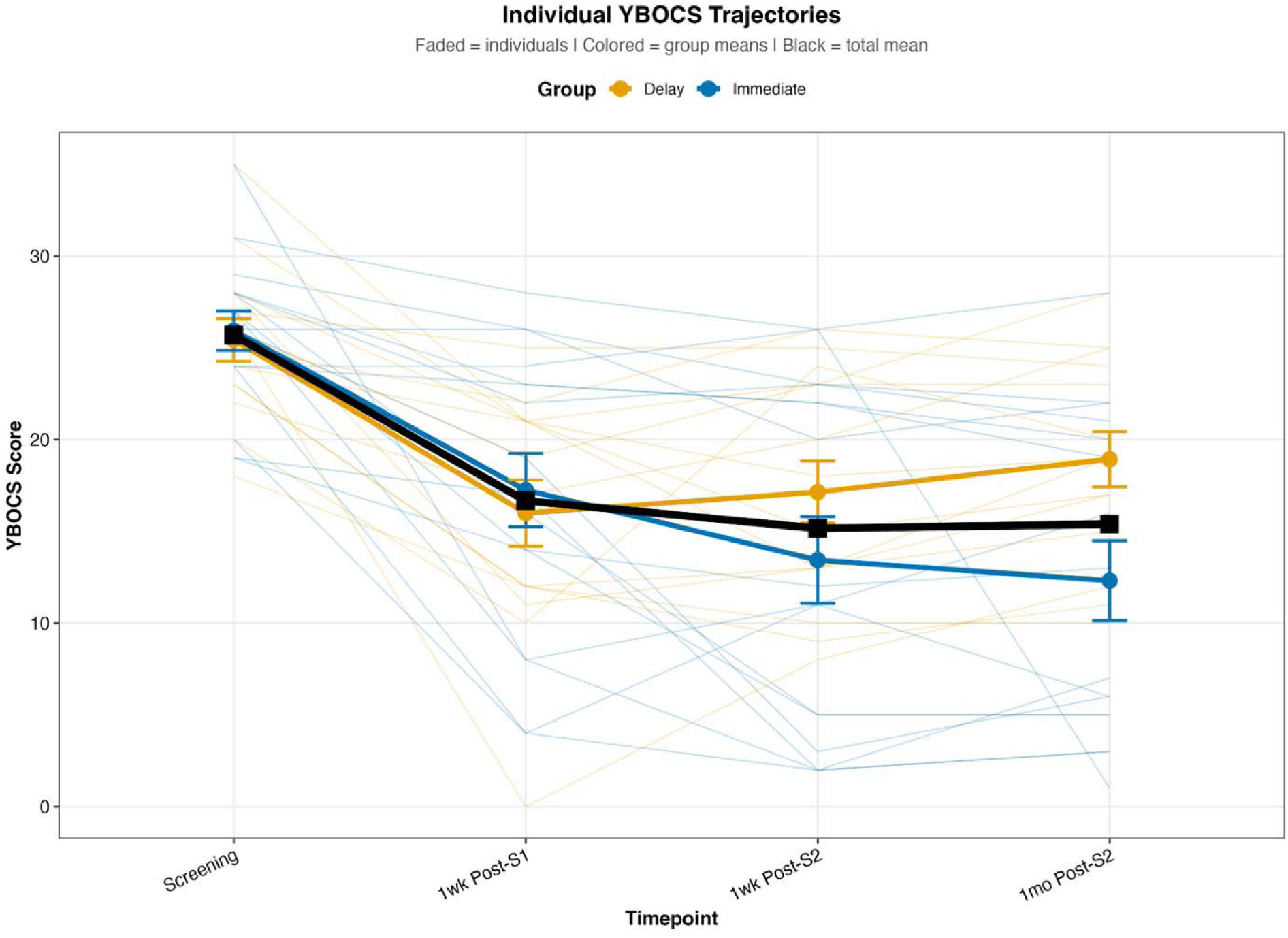
Change in Y-BOCS score before and after psilocybin treatment in all participants compared across immediate and delay groups: When compared before and after psilocybin treatment in all participants across immediate and delay groups, the average clinician administered Y-BOCS scores decreased over one month post session 2, but the pattern of decrease was slightly different across groups. Participants in waitlisted-control group post crossover into psilocybin administration showed a significant difference in their average Y-BOCS score one-month post-session 2 compared to the immediate group scores. There was a significant GroupxTime interaction (F(3,84)=4.17, p=0.008, η^2^□=0.13) for a pre/post psilocybin treatment comparison across all 30 participants for measures after each session up to one month follow-up with a main effect of time (F(3,84)=29.91, p<0.001, η^2^□=0.52) but no main effect of group (F(1,28)=1.12, p=0.300, η^2^□=0.04). Post-hoc analysis showed that Y-BOCS scores decreased one week after session 1 (Immediate: p<0.001, Cohen’s d=1.36; Delay: p<0.001; Cohen’s d=1.66) and session 2 (Immediate: p<0.001; Cohen’s d=1.7. Delay: p=0.002; Cohen’s d=1.52), as well as at one month post second dosing session (Immediate: p<0.001; Cohen’s d=1.98. Delay: p=0.004; Cohen’s d=1.29) as compared to their respective average baseline scores. However, post-hoc analysis revealed that the delay group’s Y-BOCS scores were higher than the immediate group at the one-month follow-up (p<0.05; Cohen’s d=0.95).

**eFigure 2:**
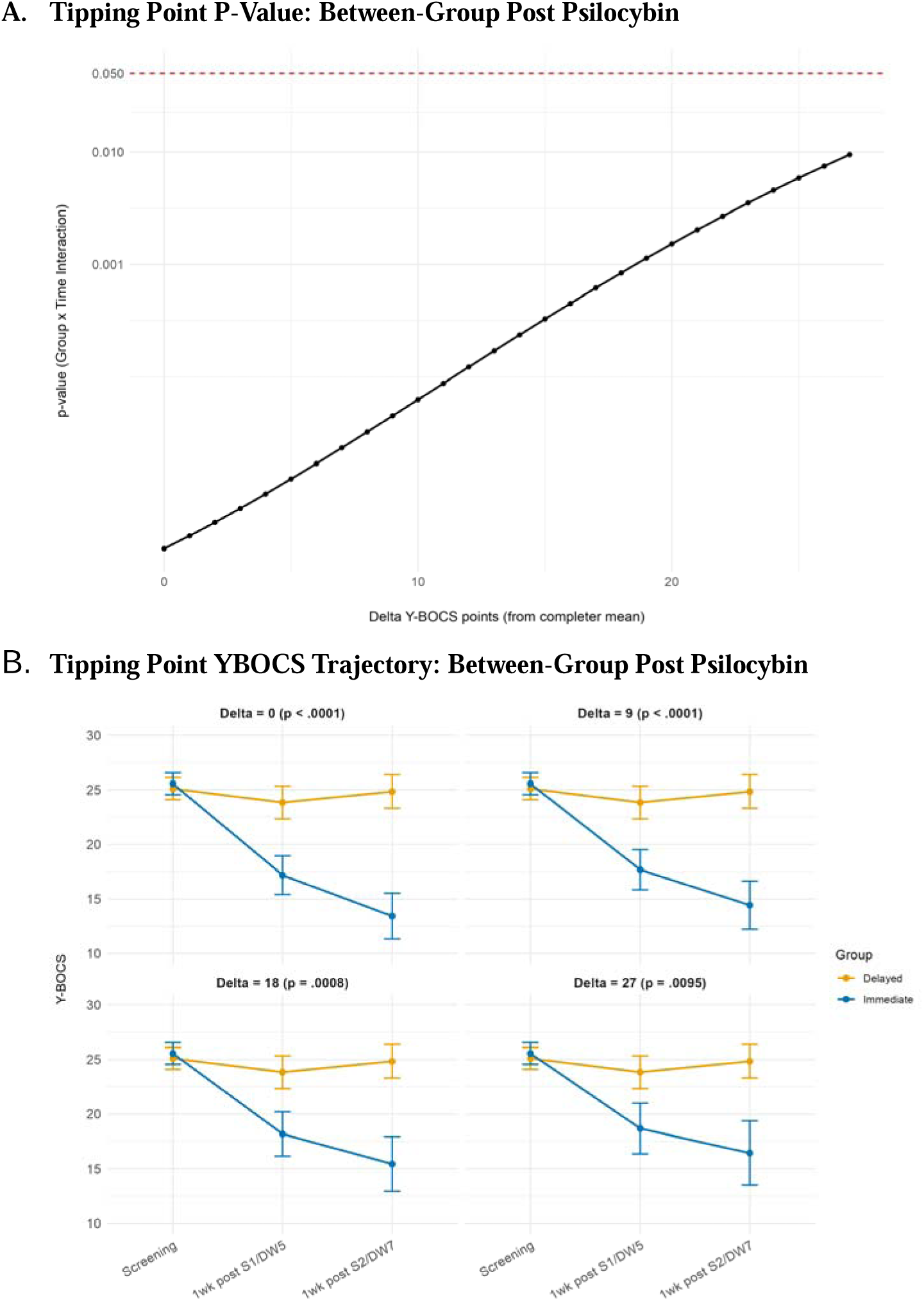
Tipping point sensitivity analysis for between-group treatment-versus-waitlist comparison. Following the treatment-versus-waitlist tipping point analysis, the Group x Time interaction remained significant across all tested delta values (delta=0: F=15.3, p<0.0001; delta=27: F=4.98, p<0.01). No tipping point was identified, indicating that the between-group difference in Y-BOCS trajectories during the treatment-versus-waitlist period was robust to missing data. Missing Y-BOCS scores for the two withdrawn participants from the immediate group were imputed using the completer mean at each timepoint, adjusted by a delta parameter representing progressively worse outcomes. The dashed red line in panel A indicates α = .05. No tipping point was identified; the group × time interaction remained significant even at delta = 27, where imputed scores reach the Y-BOCS ceiling of 40.

**eFigure 3:**
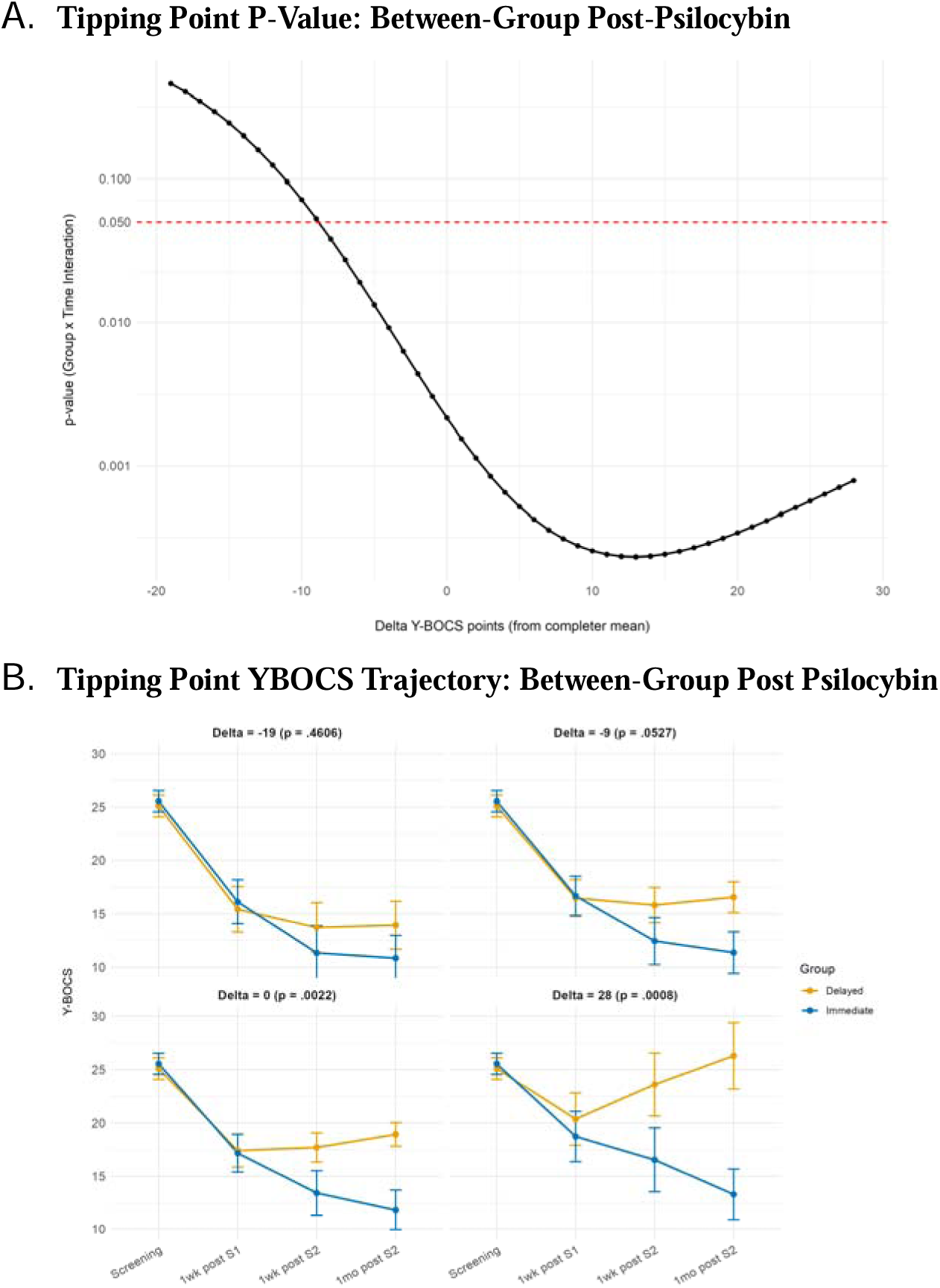
Tipping point sensitivity analysis for between-group post-psilocybin administration for all participants.. The post-psilocybin tipping point analysis showed a significant Group x Time interaction at delta=0 (F=5.20, p<0.01) and remained significant across all positive delta values through delta = 28 (F=6.02, p<0.001). The interaction became non-significant at delta = −9 (F=2.65, p=0.053), a scenario requiring withdrawn participants to have scored 9 Y-BOCS points better than completers in their respective groups to shift the results. This finding indicates that both treatment groups followed different Y-BOCS trajectories across the full study timeline even after both received psilocybin, and that this difference is not attributable to missing data. Missing Y-BOCS scores for the seven withdrawn participants were imputed using the completer mean for their respective treatment group at each timepoint, adjusted by a delta parameter. Negative delta values assume dropouts scored better than completers; positive values assume worse outcomes. The dashed red line in panel A indicates α = .05. The tipping point occurred at delta = −9, where the group × time interaction became non-significant (p = .053), requiring dropouts to have scored 9 points better than completers – an unlikely scenario.

#### eMethods 4: Self-assessed quality of life enjoyment and satisfaction, depression, and anxiety significantly change after psilocybin treatment

Difference in self-assessed scores between immediate-group and waitlist-control at time matched measures for baseline scores, 1-week post session 1 vs delay week 5, and for 1-week post session 2 vs delay week 7 is shown in eFigure 4. We find no significant interaction of Group and Time, or any main effect of time or group on Q-LES-Q (F(2,56)=2.53, p=0.088, η^2^□=0.08) and BDI-II scores (F(2,56)=0.63, p=0.53, η^2^□=0.02), but we note a significant interaction for both STAI-State (F(2,56)=3.49, p=0.037, η^2^□=0.11) and STAI-Trait (F(2,56)=7.53, p=0.001, η^2^□=0.21) anxiety measures, with a main effect of time on state anxiety measures (F(2,56)=4.91, p=0.011, η^2^□=0.15) in an expected direction.

When pre- and post-psilocybin scores were compared across all participants after delay group crossover into psilocybin intervention, self-reported mental health and well-being measures showed a general decrease following treatment relative to baseline with time. Participants completed baseline assessments to self-report quality of life enjoyment and life satisfaction, severity of depression, and anxiety before and after psilocybin sessions and followed up to one month post session 2 (eFigure 5). There was no Group x Time interaction but there was a main effect of Time on the Q-LES-Q scores (F(3,83.1)=4.48, p=0.006, partial η^2^=0.14) and BDI-II scores (F(3,83.2)=5.94, p=0.001, partial η^2^=0.18). There was an interaction of Group x Time for both State (F(3,82.3)=2.86, p=0.04, partial η^2^=0.09) and Trait (F(3,83.0)=3.49, p=0.019, partial η^2^=0.11) anxiety measures, with Trait anxiety also showing a main effect of Time (F(3,83.0)=5.33, p=0.002, partial η^2^=0.16).

**eFigure 4:**
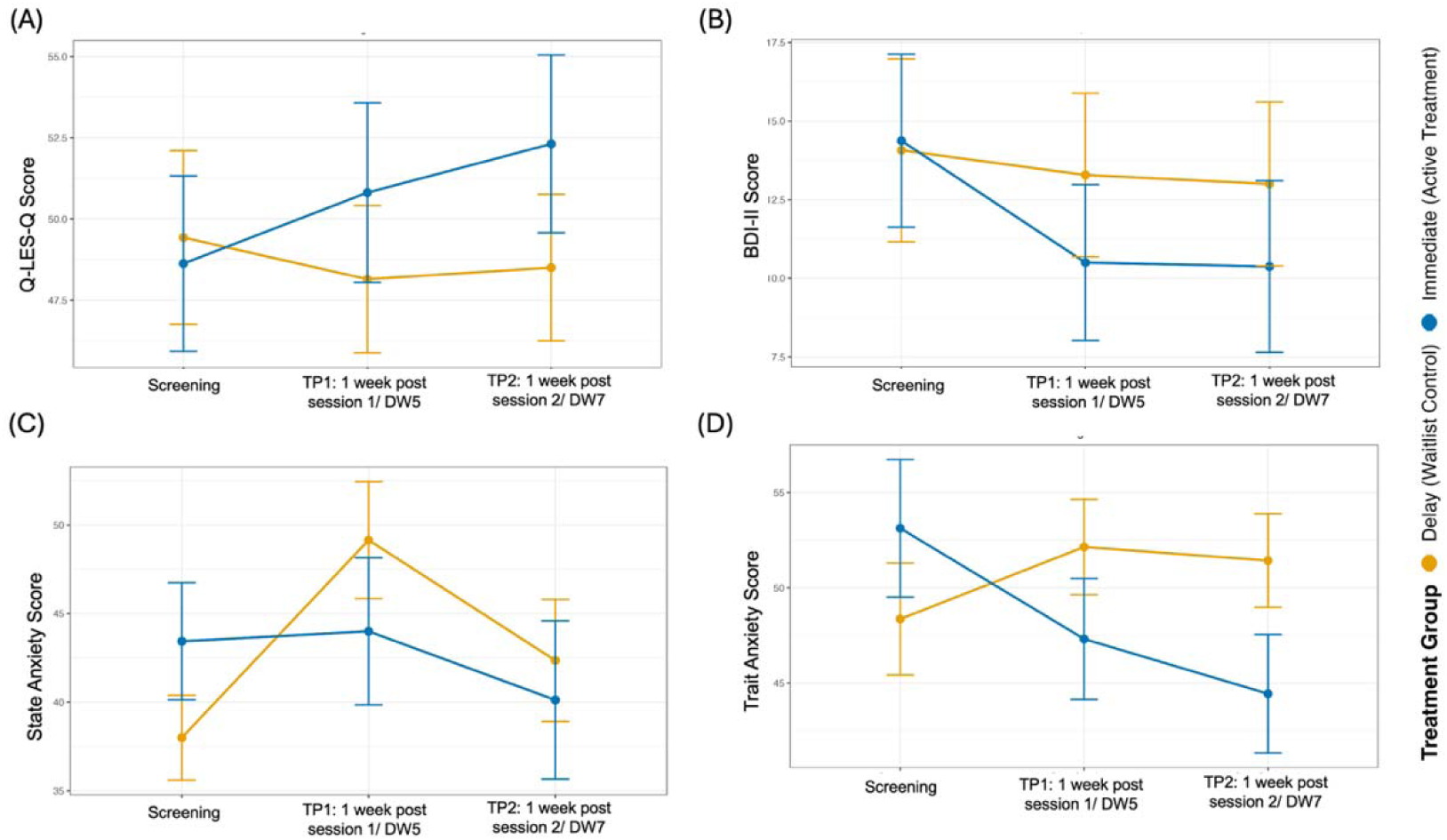
Change in self-assessment measures compared between delay and immediate groups in drug/no drug assessment arm.

**eFigure 5:**
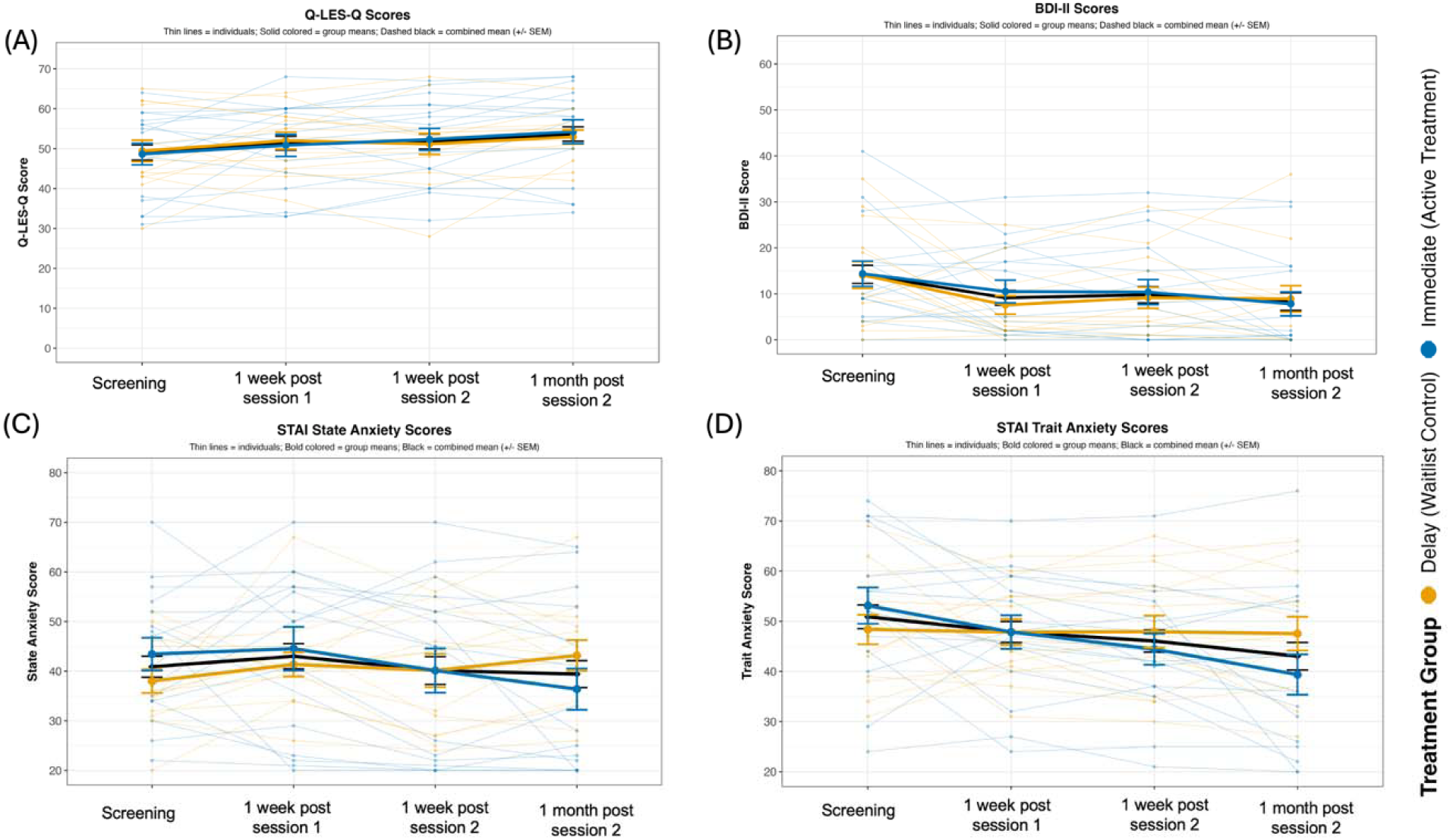
Change in self-assessment measures throughout the study course for both immediate and delay groups before and after psilocybin intervention.

## Notes

### Competing Interest Statement

SMN has consulted for Resilient Pharmaceuticals and MMS Holdings. NS serves as a paid Data and Safety Monitoring Board (DSMB) member for PT2PC, a clinical trial of psilocybin therapy for palliative care. FSB has been paid as a scientific advisor and hold stock in MindState Design Labs, LLC, and I am also a scientific advisor for Mind Pharmaceuticals. DBY has received research support from the NIH, the Heffter Research Institute, the Gracias Family Foundation, and philanthropic donors to the Johns Hopkins Center for Psychedelic and Consciousness Research. He co-directs the Hub at Oxford for Psychedelic Ethics (HOPE), which has received a small anonymous donation to support travel for underrepresented workshop participants; DBY does not personally receive financial support for this role. DBY has received consulting a fee from Soneira and educational speaking fees from the Integrative Psychiatry Institute.

### Clinical Trial

NCT05546658

### Author Declarations

IRB of Johns Hopkins School of Medicine gave ethical approval for this work under IRB00284207.

### Funding

- Funded with philanthropic support from Charlie Scheeler and Mary Ellen Pease.

